# AImmune: a new blood-based machine learning approach to improving immune profiling analysis on COVID-19 patients

**DOI:** 10.1101/2021.11.26.21266883

**Authors:** Runpeng Harris Han, Xi Tom Zhang

## Abstract

A massive number of transcriptomic profiles of blood samples from COVID-19 patients has been produced since pandemic COVID-19 begins, however, these big data from primary studies have not been well integrated by machine learning approaches. Taking advantage of modern machine learning arthrograms, we integrated and collected single cell RNA-seq (scRNA-seq) data from three independent studies, identified genes potentially available for interpretation of severity, and developed a high-performance deep learning-based deconvolution model AImmune that can predict the proportion of seven different immune cells from the bulk RNA-seq results of human peripheral mononuclear cells. This novel approach which can be used for clinical blood testing of COVID-19 on the ground that previous research shows that mRNA alternations in blood-derived PBMCs may serve as a severity indicator. Assessed on real-world data sets, the AImmune model outperformed the most recognized immune profiling model CIBERSORTx. The presented study showed the results obtained by the true scRNA-seq route can be consistently reproduced through the new approach AImmune, indicating a potential replacing the costly scRNA-seq technique for the analysis of circulating blood cells for both clinical and research purposes.

## Introduction

Severe acute respiratory syndrome coronavirus 2 (SARS-CoV-2), a novel strain in the family of the genus *Betacoronavirus*, is the pathogen that causes coronavirus disease 2019 (COVID-19) [1], which was officially declared as a global pandemic by the World Health Organization in March 2020. However, the clinical manifestations of COVID-19 show great heterogeneity, from asymptomatic infections or a moderate upper respiratory tract infection to severe viral pneumonia some of which may cause cytokine storm and cause the mortality of the patient [2, 3]. Early studies reported that hyperinflammatory responses to SARS-CoV-2 infections are related to disease severity and mortality, and the monitoring and prediction of hyperinflammatory responses could emerge as methods to detect high-risk patients and give specific treatments based on their clinical characteristics. A variety of previous studies [4, 5] have reported several aberrant immune phenomena relating to disease severity, including lymphopenia [6], Monocyte-Macrophage abnormalities [7], impaired interferon activity [8], and cytokine storms [9]. Indeed, these studies do contribute to revealing several potential immunomodulatory agents in attenuating the overactivated immune response to SARS-CoV-2 and obtain better patient outcomes.

With the development of sequencing technologies, a large amount of scRNA-seq data has been generated in the study on SARS-CoV-2, yet these data from independent studies are not well utilized except for integration and meta-analysis, while machine learning methods, which can learn and abstract pattern and connections from a large amount of data, ideally suited to reanalyze these data and thus provide us with new understandings of the development of COVID-19. The composition of immune cells in the blood, especially the changes in the ratio of each lymphocyte subpopulation, can be used as biomarkers for diagnosis and severity prediction of COVID-19 [10-14]. However, the current tools available for clinical testing of immune cell composition, such as flow cytometry and single-cell sequencing, are still pricy, which thwart immune profiling from being extensively used. In the present study, we integrated scRNA-seq from 3 different studies, validated the results from previous independent studies with the help of machine learning technologies, including the traditional tree-based techniques and the new deep learning techniques, we screened molecular biomarkers that could be used to indicate disease processes in much higher throughput than manual selection. Here we report the application of a novel deconvolution model AImmune that can predict the proportion of seven different immune cells utilizing bulk RNA-seq results of human peripheral mononuclear cells (PBMCs). Assessed on real data sets, the model outperformed the well-recognized immune profiling models CIBERSORT and CIBERSORTx [15, 16]. Through the model, the results obtained by the true scRNA-seq route can be well reproduced, thus the AImmune model has the potential in replacing the costly scRNA-seq technique for the analysis of immune cell composition for both clinical and research purposes.

## Result

### PBMC-based molecular features identified in COVID-19 subgroups versus healthy subjects

In order to summarize the differentially expressed genes (DEGs) in peripheral blood mononuclear cells (PBMCs) from COVID-19 patients, a multi-center scRNA-seq cohort was built by selective COVID-19 datasets collected from Gene Expression Omnibus (GEO) (**Figure 1** & **Table 1**). The combined cohort is composed of three independent datasets (GSE150728, GSE163668, and GSE166489), covering two COVID-19 patient subgroups defined by severity (Severe and Moderate) as compared with Healthy donor group (Healthy). In total 23 subjects (25,9339 individual cell profiles) were included in the final cohort after data preprocessing (merging, quality control, and batch correction), including 23 healthy subjects (133,055 cells), 10 moderate cases (53,479 cells) and 12 severe cases (72,805 cells) (**Table 1**). Standard bioinformatics analysis identified 1883, 1382 and 236 DEGs (adjusted P-value < 0.05 and Log2 fold-change >=1) in comparison of Healthy vs. Moderate, Healthy vs Severe and Moderate vs Severe, respectively (**Figure 2A**).

**Table 1.** Demographics and GEO accession ID of enrolled studies.

| GEO ID | Severity Group | Subject# | Gender (F:M) | Age (y) | Race | Cell# |
| --- | --- | --- | --- | --- | --- | --- |
| GSE150728 | Healthy | 6 | 2:4 | Avg. = 44.7 | White: 5<br>Asian: 1 | 15054 |
| GSE166489 | Healthy | 7 | 2:5 | Pediatric: 4<br>Adult: 3 | Not provided | 45269 |
| GSE166489 | Moderate | 3 | 3:0 | Pediatric: 3 | Hispanic: 2<br>Latino: 1 | 30757 |
| GSE166489 | Severe | 5 | 0:5 | Pediatric: 5 | Not provided | 29391 |
| GSE163668 | Healthy | 10 | 4:6 | Avg. = 38.7 | Not provided | 72732 |
| GSE163668 | Moderate | 7 (including 1<br>pooled sample) | 2:4 | Avg. = 59.3 | Not provided | 22722 |
| GSE163668 | Severe | 7 (including 3<br>pooled samples) | 2:2 | Avg. = 52.1 | Not provided | 43414 |
| **Total** |  | **45** | **15:26** |  |  | **259339** |

**Figure 1.**
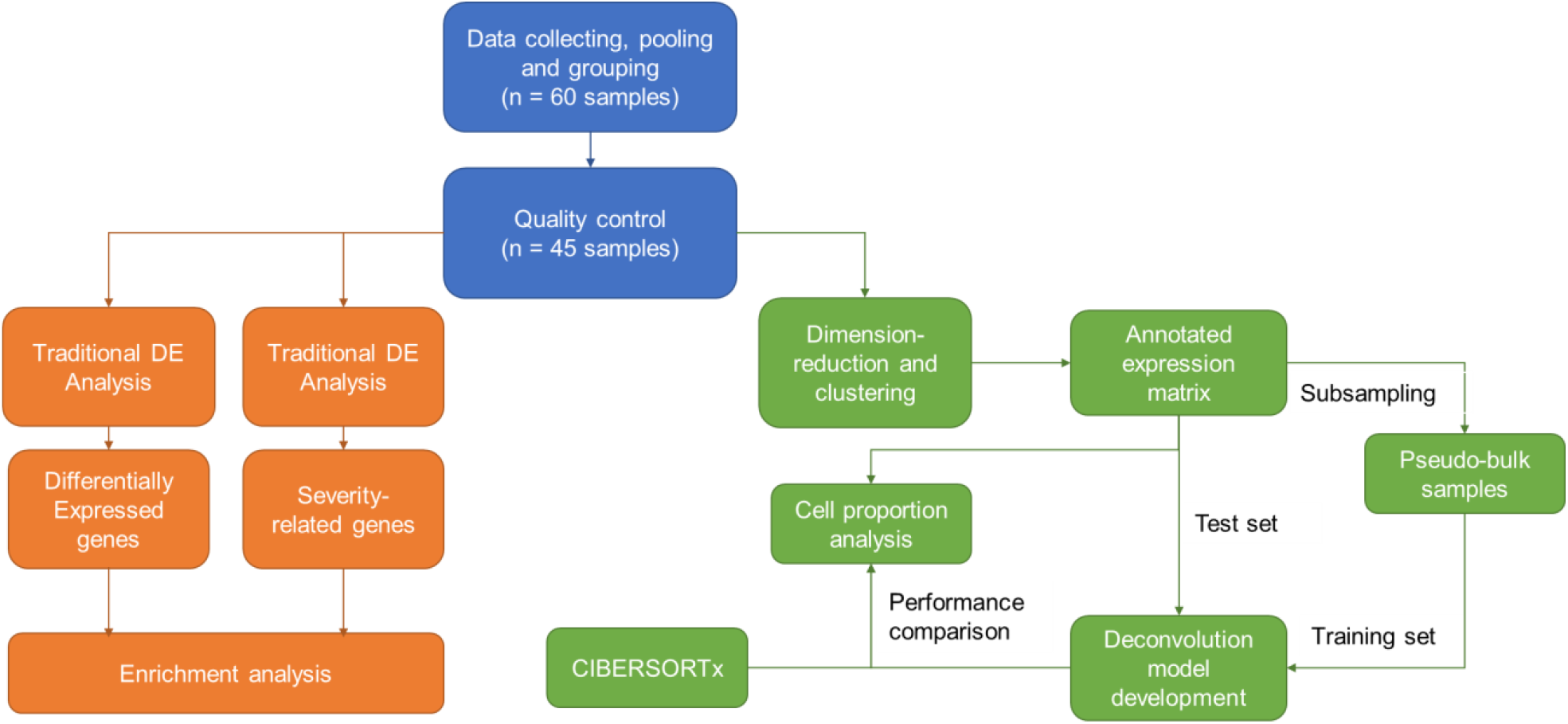
Flowchart of the present study. DE: differential expression; ML: machine learning.

**Figure 2.**
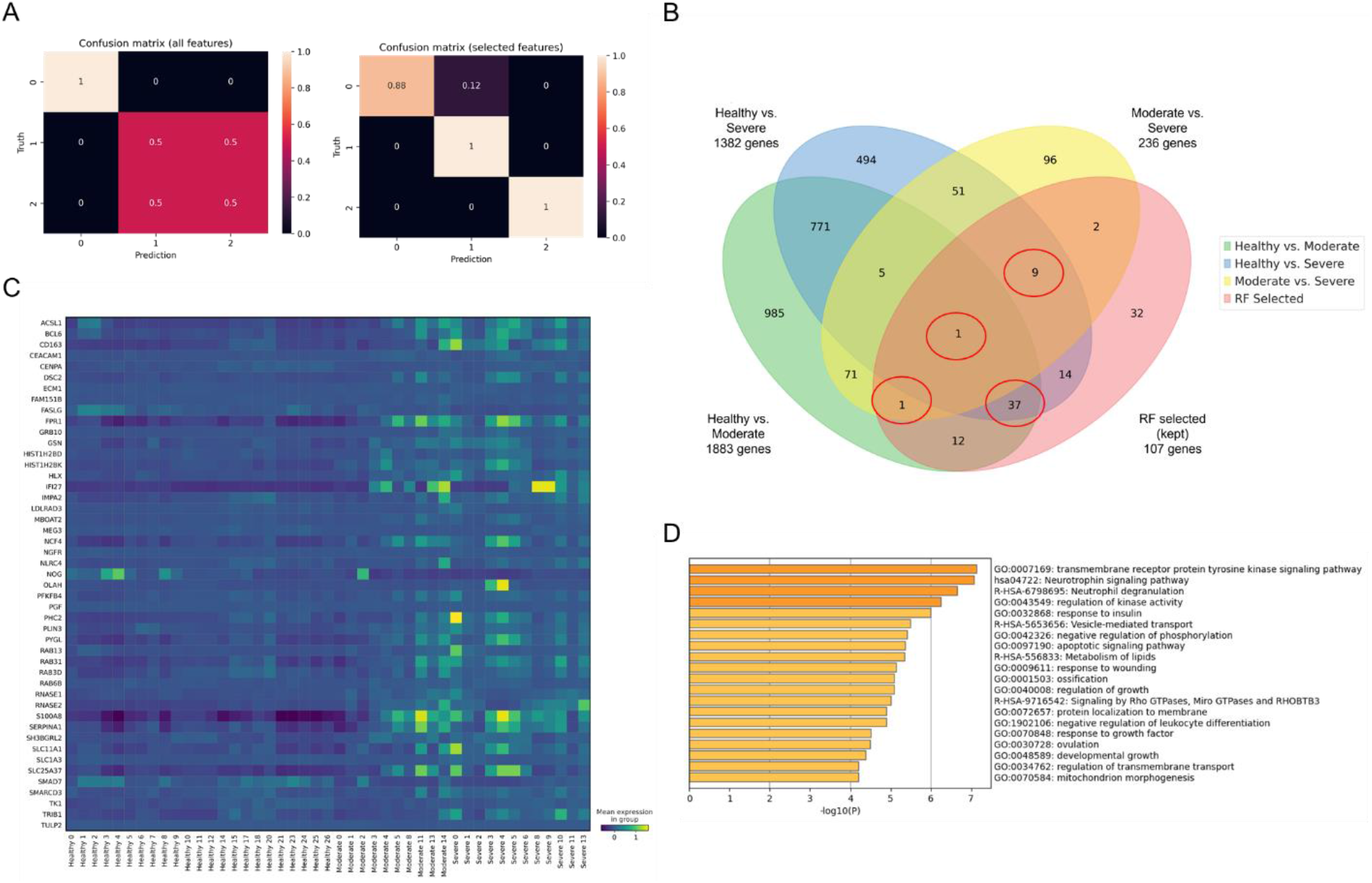
Random Forests algorithm identified PBMC-based gene predictors with improved performance in predicting COVID-19 clinical severity. A) Confusion matrix of predicting results by random forest models using all gene predictors (left) and 107 selected gene predictors (right). Numbers 0, 1 and 2 indicate Healthy, Moderate and Severe subgroup. B) Venn diagram showing the overlaying of DEGs obtained from traditional different gene analysis and selected gene predictors (top one third of relative importance, n=107) identified by Random Forests-based classification. Red cycles indicate DEGs showed in at least traditional analysis comparisons and also identified as gene predictors by Random Forests algorithm (total n = 48). C) Matrix plot depicting average expression of 48 overlaying genes identified by Venn diagram (B). D) Gene Ontology enrichment analysis using 107 selected gene predictors identified by Random Forests model.

### PBMC-based gene predictors selected and optimized by Random Forests algorithm

Random Forests (RF) is a popular tree-based machine learning algorithm that has been successfully applied in genetic data. Comparing to other machine learning or deep learning algorithm, RF as a classifying approach is well suited to small sample studies. Here using the gene expression profile as predictors and the severity status as the outcomes, a non-parametric RF model was built relating the predictors to the outcome (sample size = 45) with a predicting accuracy of 0.833 (**Figure 2A, left**). With the hyper-parameters tested and optimized (max depth = 10, split = Gini impurity) to ensure the best predicting performance, 107 DEGs (top one third of relative importance) were identified from in total 17206 genes as a panel of key PBMC-based molecular features that can distinguish individual COVID-19 subgroups from Healthy group (**Figure 2B**). Replacing the overall gene predictors by the top ranked predictors could improve the prediction accuracy by 8.3% (from 0.833 to 0.916) (**Figure 2A, right**). Most of them showed higher expression in Severe cases and lower expression in Healthy subjects (**Figure 2C**). Notably, 70.1% (n = 75) of these marker genes were overlaid with DEGs from at least one list identified above through traditional analyses (**Figure 2B**). Interestingly, removing the screening restriction (relative importance > 0) would yield a bigger size of molecular features (n = 321), while the rate of overlaying marker genes in this predictor panel dropped down to 42.7% (n = 137) (**Supplementary Figure S1**). Gene Ontology enrichment results showed that degranulation of neutrophils was a major immune response against COVID-19 since related signaling pathway was enriched in all four DEG lists as showed above (**Figure 2D & Supplementary Figure S2**). In contrast, Moderate subgroup displayed additional layer of immune regulation or activation, especially cytokine regulation, as compared to Severe cases.

### The compositions of key immune cells were related with COVID-19 severity

Next we sought to profile the composition of immune cell subsets in COVID-19 patients and compare them with healthy subjects and between subgroups. In order to make this analysis through bulk cell RNA-seq data collected from PBMCs, we developed AImmune, a deep neural network (DNN) model trained by the COVID-19 scRNA-seq dataset that can deconvolute 6 cell subsets (CD4+ T cells, CD8+ T cells, Monocytes, NK cells, B cells, DC cells) and platelets. Firstly, the gene expression profiles from entire cell samples were clustered into cell subset groups as labelled by pre-defined cell subject markers (**Figure 3A & 3B**). The pre-defined markers were previously collected and validated by single cell analysis toolkit Scanpy. The expression status of these pre-defined marker genes in our cell samples were illustrated (**Figure 3C**). The un-supervised clustering provided by AImmune allowed closer quantitative analysis of immune cell subset (**Figure 3D**). Interestingly, lower cell numbers were estimated in the T lymphocytes, NK cells and DC cells in COVID-19 Moderate subgroup and even lower in Severe subgroup (all P<0.05). On the other hand, monocytes were significantly more abundant in COVID-19 cases (all p<0.05) than in healthy controls.

**Figure 3.**
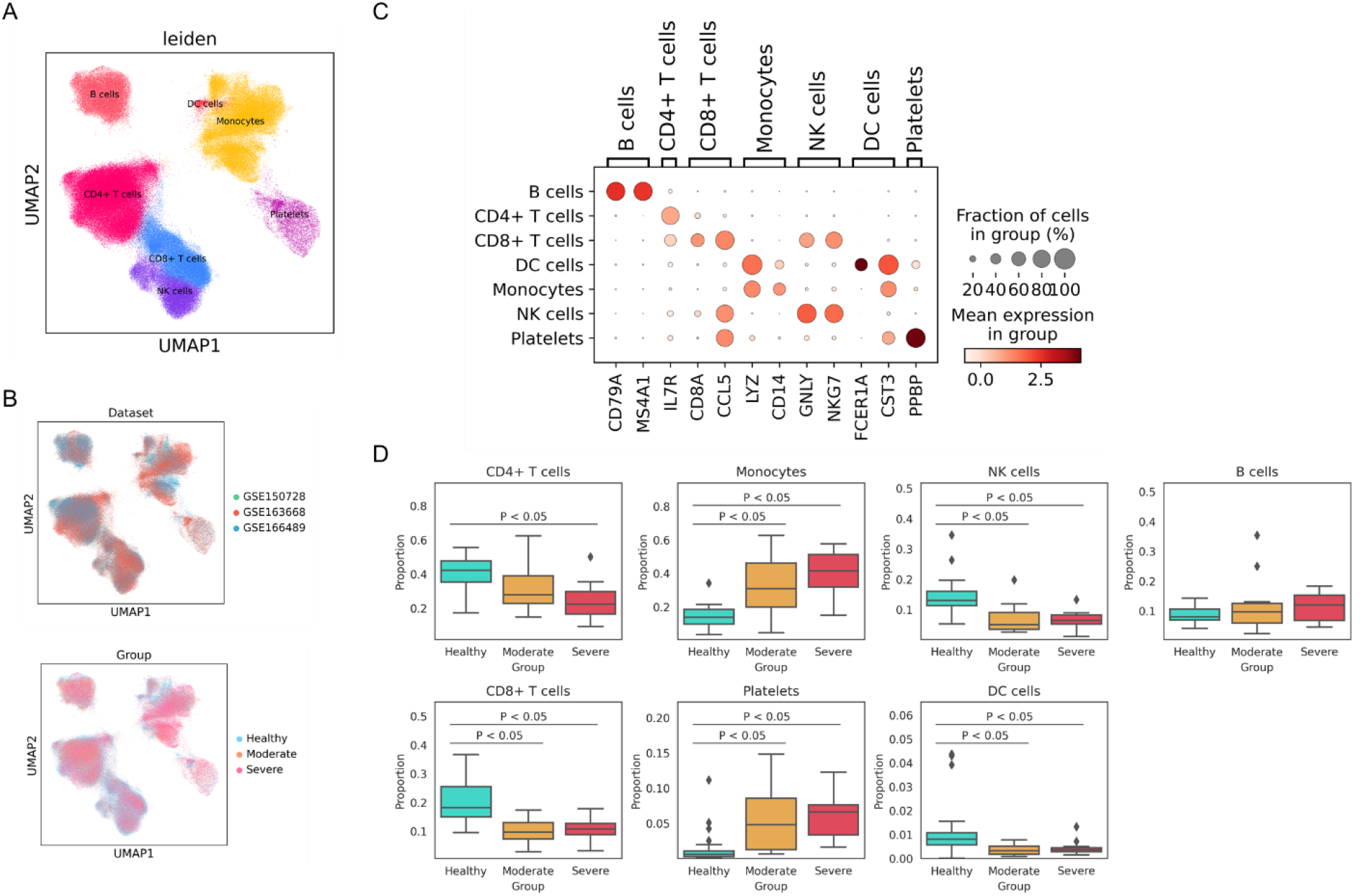
Immune cell compositions were consistently changed between COVID-19 severity subgroups and healthy subjects. A) UMAP embedding of the total cells in the combined dataset (n= 259,339 cells) colored by Leiden algorithm-generated clusters, marked by manual cell type annotation using predefined markers (see panel C) as references. B) UMAP dimensionality reduction embedding of the total cells marked by dataset sources (upper) and severity group (lower). C) Dot plot showing the average expression level (color scale) and expressing rate (size scale) of predefined marker gene in cell type. D) Box plot comparing cell compositions of each cell subset in each colored annotated group. Two-sided P values by the Welch’s t-test less than 0.05 are shown.

### AImmune showed reliable performance in predicting immune cell compositions in COVID-19 PBMCs

Our hypothesis is this new approach AImmune would yield comparable prediction performance with the existing immune profiling methods. Unlike the widely recognized immune cell deconvolution model – CIBERSORTx, AImmune does not require signature matrixes and uses DNN rather than support vector machines (SVMs) as regression model, which were been proven in previous studies by generating a large amount of pseudo-bulk data to train, DNNs can achieve a decent performance [17]. Although previous studies have argued that because DNNs can automatically select features by assigning weights and adding L1 regularization, no dedicated feature selection work is needed, we conducted a tree-based feature selection before using datasets for training. By testing on a three-layer DNN built by three different feature-selection strategies, our result showed all variables or ML/RF selected features worked better than highly variable features (**Figure 4A**). We then compared the performance of CIBERSORTx and AImmune on the 45 datasets as mentioned above. It turned out that AImmune approach showed a better predicting performance as the average mean squared error (MSE) across all cell subsets is much lower in AImmune than CIBERSORTx (0.0079 versus 0.016) (**Figure 4B)**. The average cell abundance levels and Pearson correlation of individual cell composition values were compared between ground truth and predictions made by AImmune and CIBERSORTx (**Figure 4C & 4D)**. Our results demonstrated the predicted value generated by AImmune is highly consistent and correlated with ground truth values, which seems even better than prediction made by CIBERSORTx. Together, we concluded that AImmune is reliable in predicting compositions of immune cell subsets.

**Figure 4.**
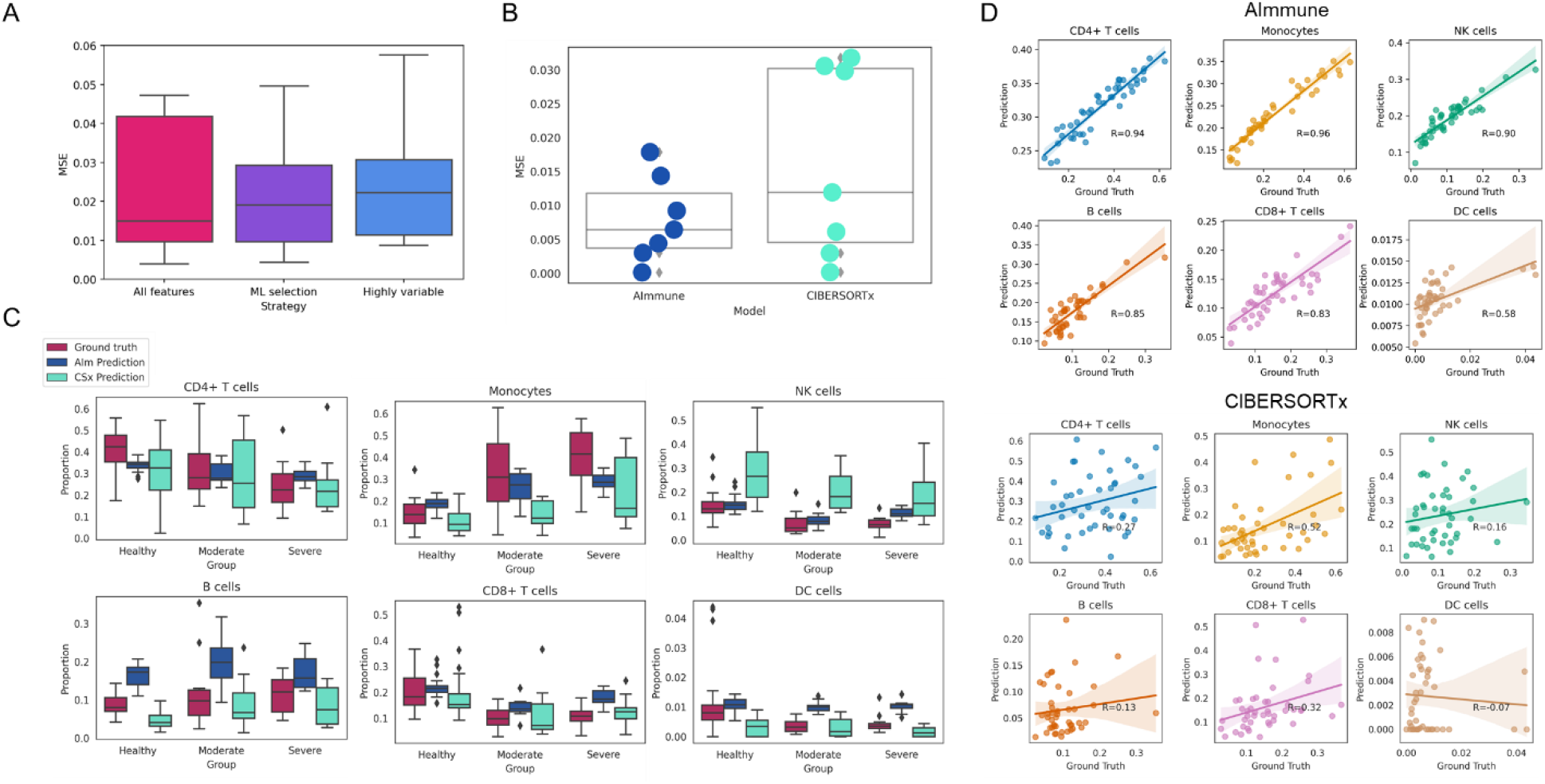
AImmune produced reliable prediction on immune cell compositions. A) Boxplots of the prediction Mean-Square Error (MSE) of AImmune models (3-layer fully-connected neural networks with same number of nodes) built three different feature-selection strategies. B) Boxplots and scatter plots of the prediction MSE of predictions from AImmune and CIBERSORTx for 6 cell types and palatae. C) Boxplots comparing the ground truth composition levels with values predicted by AImmune and CIBERSORTx for 6 cell subsets. Prediction from CIBERSORTx for Platelets is excluded since the LM22 signature matrix does not include item of Platelet. D) Scatterplots of ground truth (x-axis) and predicted values (y-axis) generated from AImmune (up) and CIBERSORTx (down) for 6 cell types. Pearson’s Pearson correlation coefficient values are given as inserts.

## Discussion

Although it has been reported previously, the present study investigated and confirmed the immune cells abundances are differently observed in COVID-19, which is broadly supported by previous studies[18-22]. This study also reported a novel computational approach enables blood cell-based immune cell profiling in both clinical and research settings at a much lower cost than using scRNA-seq methods, which in turn allows for larger scale, broader applications, and studies.

Through our integrated, machine learning-facilitated analysis of three published scRNA-seq datasets, new information which may shed lights on immunopathology of COVID-19, especially on what genes may serve as potential biomarkers predicting disease progression and severity was extracted from extent data, a novel deconvolution model was developed, and last but not least, previous findings from each independent study were validated. Integrated study confirms some of previous findings such as COVID-19 patients do have a lower proportion of T cells and NK cells in their PBMCs compared to healthy donors, but there are some inconsistencies, for example, one study from which we obtained data found that there are significant differences in B cell proportions between patients with severe COVID-19[23] while our integrated study and another independent study[24] perceived opposite results. These inconsistencies suggest that while single-cell sequencing is so expensive to conduct that sample sizes are usually smaller when conducting comparative studies with it and there are still pronounced batch effect, though it can provide many great unprecedented insights, many of the results still needs validation. In addition to immune cells, it is important to understand the role that some crucial genes and the signaling pathways they are involved in play in COVID-19 disease, unlike traditional statistical tests that can only detect genes by differential expression and always produce thousands genes, supervised machine learning methods focus on genes that influence the decision making on whether it is expression spectrum of healthy people, patients with mild COVID-19 or patients with severe COVID-19, through which we obtained 360 informative genes. Some of them are reported as potential biomarkers or important pathological genes, for example S100A8 and the calprotectin production pathway it is involved in was reported to serve as biomarkers distinguishing between mild and severe disease states[25, 26], FPR1 was reported to be a candidate surface and druggable targets FPR1 for drug delivery[27], defective Bcl-6+ TFH cell generation and dysregulated humoral immune induction early in COVID-19 disease was identified and reported as well[28], these findings along with others[29-32] validates that our gene lists do contains meaningful genes. However, there are genes in the list which have not been studies in depth, such as SLC25A37, FAM151B, PYGL, and TKT. Through studying these genes, it is possible to discover new biomarkers or drug targets. Our research also shows that analysis using machine learning methods can be used as a complement screening tool to traditional DE analysis, to discover genes that are worth investigating. In addition to helping the discovery of novel potential biomarkers and drug targets, we developed AImmuneusing our integrated data to help utilize a current known biomarker of COVID-19: Percentage of immune cells in peripheral blood. We believe that AImmune, based on its high prediction accuracy, is expected to replace expensive flow cytometry and ScRNA-seq, increasing the accessibility of immune profiling in clinical applications. In summary, our study shows the application and value of machine learning techniques in utilizing sequencing data, and we believe that the introduction of machine learning techniques into biomedical research will bring more benefits in the future.

Unlike statistical DE analysis that did not find specific pathways (supp) that could be used to differentiate patients with severe symptoms from those moderate symptoms, some very interesting pathways were found by machine learning methods, such as tyrosine kinase signaling pathway, response to insulin, and lipid metabolism, which were also reported in some studies[33-35]. This suggests that machine learning methods, especially tree-based methods, can be used as a high-throughput discovery tool in the discovery of biomarkers or drug targets for some complex diseases.

Definitely, this research has limitations: 1) Though compared with each single study where the dataset is obtained, our integrated study could offer a more general understanding, however, since the subsequent machine learning process requires datasets with severity labels, this greatly limits our optional data sources, resulting in a final cohort of 45 samples, which still not enough to provide very generalized results. 2) Since the datasets comes from different studies which means there are great consistencies in sample acquisition, processing and sequencing procedures, these differences in technical details combined with their own batch effects result in our inability to make too fine a distinction between cell subpopulations, wasting the advantages of single-cell sequencing techniques, while one of which is to provide a finer understanding of immune cell subpopulations than currently exist [36], and COVID-19 patients do have changes in some specific subpopulations of immune cells [37]. 3) Although AImmune has been tested to have high prediction accuracy and better performance than the best model of its kind, CIBERSORTx, its prediction on DC cells is relatively poor (R^2^ = 0.58, while R^2^ of predictions on other cell types are all above 0.80), and interestingly, the same situation can be observed on CIBERSORTx poor (R^2^ = -0.07, while R^2^ of predictions on other cell types are all above 0.10), we hypothesize that this is because DC cells and Monocytes have close expression profiles and DC cells do not have many marker genes with sufficient specificity. Therefore, some of the results in this experiment still need to be validated by larger-scaled and more in-depth studies and for our model, improvements and refinements are still needed to be able to predict more immune cell subpopulations with higher accuracy.

## Methods

### 1. Datasets and preprocessing scRNA-seq datasets

The protocol was approved by the local Ethics Committee and the Institutional Review Board of Norwest University (approval number: 200402001) and all patients provided written informed consent. The human PBMC scRNA-seq datasets were downloaded from Gene Expression Omnibus (GEO) with accession IDs GSE150728, GSE166489, and GSE163668, respectively (**Figure 1**). After loading and integrating these three datasets, the aggregate dataset was processed using the Python package Scanpy (v. 1.2.2) [38], following Scanpy’s reimplementation of Seurat’s clustering tutorial for 3k PBMCs from 10x Genomics workflow[39]. First, the raw count matrix was filtered for cells with less than two hundred detected genes, with too much mitochondrial gene expression (accounted for more than 5% of total counts), or too many total counts (more than 2500 genes by counts), and genes that not sequenced in all three datasets. Gene expression was normalized to ten thousand total counts and logarithm transformed through the means of function “normalize_per_cell” and “log1p” in the Scanpy package so that counts became comparable among cells. The filtered and normalized aggregate dataset was saved for subsequent pseudo-bulk datasets generation. Then the dimension of the aggregate data was reduced through the means of Principle Component Analysis(PCA) using only expression of highly variable genes which was determined through the “highly_variable_genes” function. Before using these principal components for clustering analysis, the Harmony algorithm[40] was used to correct batch effects. Because Harmony is a batch effect correction algorithm based on PCA results, the Harmony algorithm processed PCA results are used instead of the original PCA results in the neighborhood computing required for the next clustering analysis. All cells were clustered using the Leiden algorithm after that. The markers used to annotate the cell types are as follows: Interleukin-7 receptor (IL7R) was taken as a marker for CD4+ T cells, LYZ and CD14 for monocytes, CD79A and MS4A1 for B cells, GNLY and NKG7 for natural killer (NK) cells, FCER1A and CST3 for dendritic (DC) cells, CD8A and CD8B for CD8+ T cells, and PPBP as the marker Platelets.

### 2. Generating pseudo-bulk RNA-seq samples from scRNA-seq data

It is widely known that DNN requires large amounts of training data with labels to achieve an ideal performance, and on this occasion, bulk RNA-seq results with known cell compositions, which makes the generation of pseudo-bulk RNA-seq samples is a crucial step in developing Aimmune 2.0 Model. To attain this, we subsampled cells with annotated labels from the aggregate dataset which was not normalized or logarithm transformed randomly, summed the expression linearly as input, and counted the fractions of certain cell types as labels. Using this procedure, we generated five thousand pseudo-bulk samples for training in total.

### 3. Preprocessing of the input data

To make the input data more suitable for machine learning algorithms, the input data should be further transformed. One major step is to make sure that features from any sample should be at the same scale, to attain this, all the pseudo-bulk samples were the first logarithm transformed, and then using MinMaxScaler from sklearn package, these transformed samples were scaled to the range [0,1]. The Min-max scaling process is defined as follow:

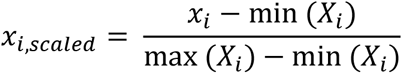

where *x*_*i,scaled*_ is the log1p expression value of gene *x* in sample *i*, while *X*_*i*_ is the vector of log1p expression values for all genes of sample *i*. Min(*X*_*i*_) is the minimum gene expression value in vector *X*_*i*_, while max(*X*_*i*_) is the maximum gene expression in vector *X*_*i*_.

### 4. Feature Selection

The raw scRNA-seq data includes the expression information of nearly 20,000 genes, yet previous researches suggest that expression information of hundreds of genes was enough for predicting the fraction of certain cells in a tissue[15]. Although DNNs can automatically learn the importance (weights) of certain features, using input data with too much irrelevant information may still affect the performance of the model. But simply using features that are involved in published signature matrixes may waste the computing power of deep neural networks since these signature matrixes are used for Support Vector Machines (SVMs), which are fairly simple models compared to DNNs, so proper feature selection methods is required. After testing a few methods including Lasso regression, SVM-based recursive feature elimination, random forests (RFs) was selected due to their ability to fit nonlinear relationships well and does not require a large amount of data. By fitting an RF regressor to the training data, the impurity-based importance of each feature was calculated, and only features with importance of more than 0.1 are kept for training.

### 5. Model Training

After the preprocessing of the data, DNNs that can predict the fraction of different cell types are trained. DNNS with three layers are first trained using three different training strategies and the mean square error on the validation set is used to indicate the model performance : training with all the features, training with highly variable features, or training with selected, cell-type-specific features, it turns out that models trained using the third strategy have the best performance, which verifies our previous hypothesis that proper feature selection matters.

After that, for predicting a single cell type, a model was optimized individually by tuning hyperparameters. To prevent overfitting, L2 regularization is introduced for each layer of all models and an early stop method with varying patience indices is introduced in the training. The L2 loss is defined as following equation:

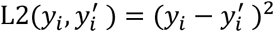

where *y*_*i*_ is the of ground-truth fractions of a certain cell type in sample *i* while *y*^*′*^ is the corresponding prediction value.

### 6. CIBERSORTxx

CIBERSORTxx (CSx) is an updated version of CIBERSORTx (CS), for deconvolution via CIBERSORTxx, we used the LM22 signature matrix, by means of which proportions of 22 leukocyte subsets can be predicted. Since the LM22 signature matrix deconvolves cell types at a more meticulous scale than what was used for this study, prediction results were added together, and since the LM22 signature matrix does not contain Platelets in its columns, so only the prediction of proportion of other 6 cell types were compared and presented.

### 7. DE Analysis

Bulk expression data was firstly normalized by the calcNormFacors function which was based on TMM algorithm in R package edgeR. Differentially expressed genes (DEGs) was then searched using by one-to-one comparison between different groups (Healthy vs. Moderate, Healthy vs. Severe, Moderate vs. Severe, respectively) and only genes with Benjamini & Hochberg (BH) adjusted P-value < 0.05 and Log2 fold-change (L2FC) >=1 were kept for subsequent analysis. All tests and comparisons are done using the built-in functions of the R package edgeR.

## Data Availability

All data produced are available online at https://github.com/HarrisHan2000/AImmune3.0, and https://github.com/HarrisHan2000/COVID_Project.

https://github.com/HarrisHan2000/COVID_Project

## Data availability

The source code for AImmune is available at https://github.com/HarrisHan2000/AImmune3.0, and the source code for other parts of this study and lists of selected genes are available at https://github.com/HarrisHan2000/COVID_Project.

## Author contributions

RHH collected and downloaded the published the data, analyzed the combined datasets, illustrated the results and wrote the manuscript. XTZ designed the research flow, reviewed and revised the manuscript and supervised all aspects of the study.

## Acknowledgments

We thank all participants of the original studies and their authors who reported and published the data in GEO.

