## Supplementary material for "AImmune: a new blood-based machine learning approach to improving immune profiling analysis on COVID-19 patients": Figure S1

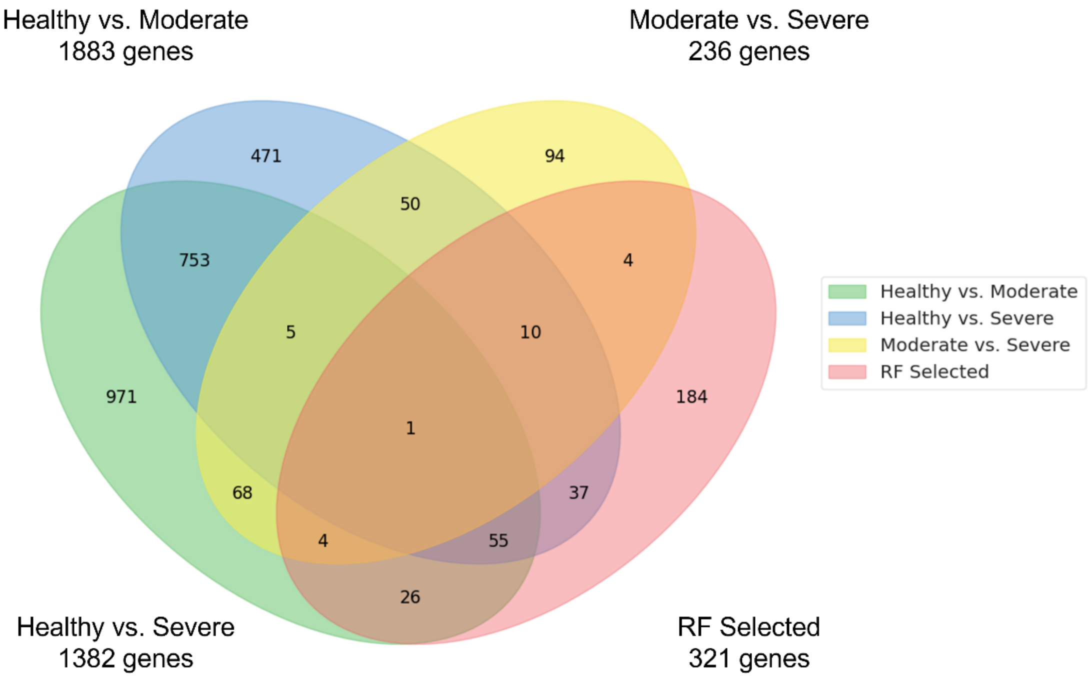


Figure S1. Venn diagram showing the overlaying of DEGs obtained from traditional different gene analysis and all gene
