## Supplementary material for "AImmune: a new blood-based machine learning approach to improving immune profiling analysis on COVID-19 patients": Figure S2

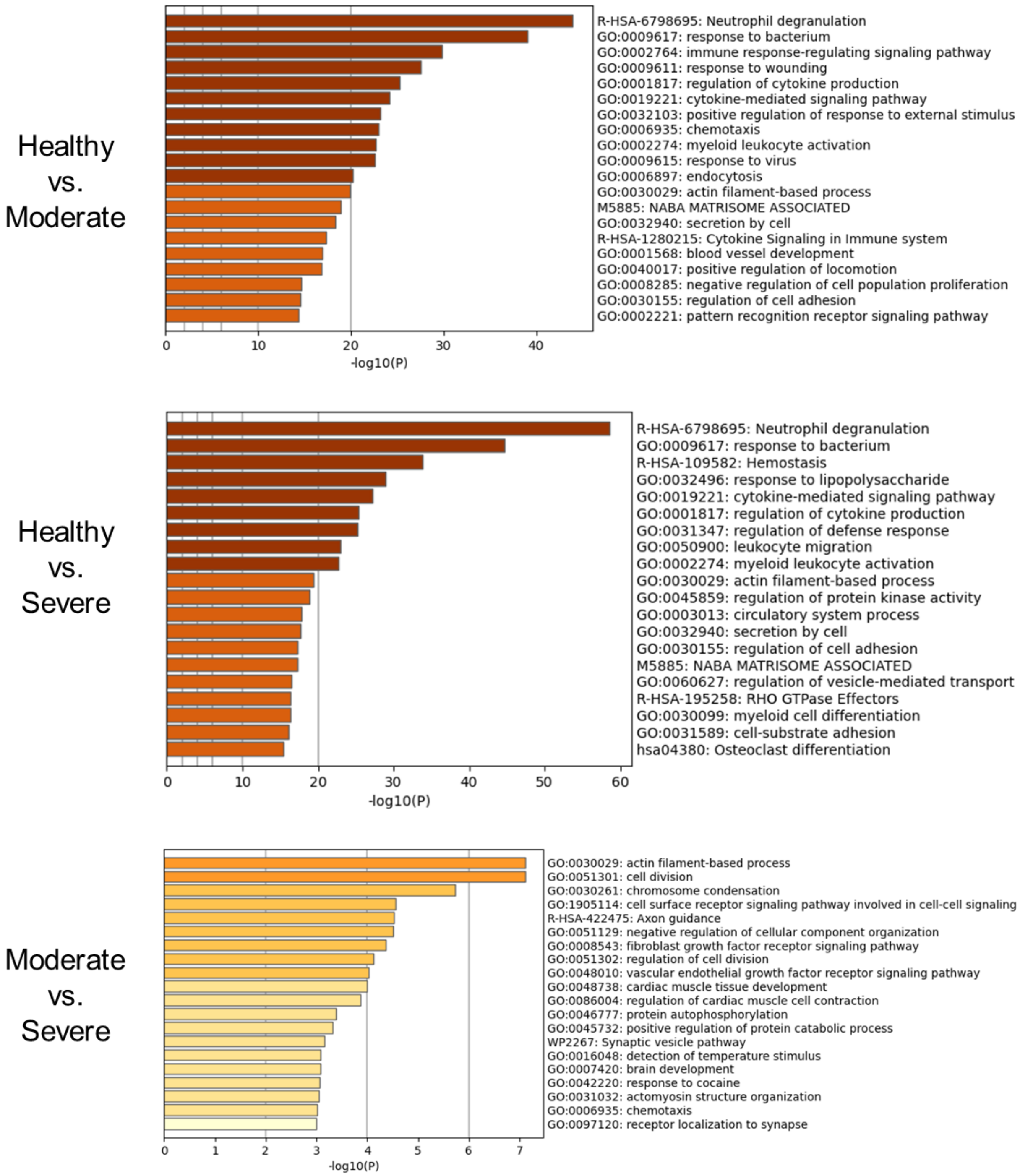


Figure S2. Gene Ontology enrichment analysis using 107 selected gene predictors identified by Random Forests algorithm.
