## Supplementary material for "AImmune: a new blood-based machine learning approach to improving immune profiling analysis on COVID-19 patients": Figure S3

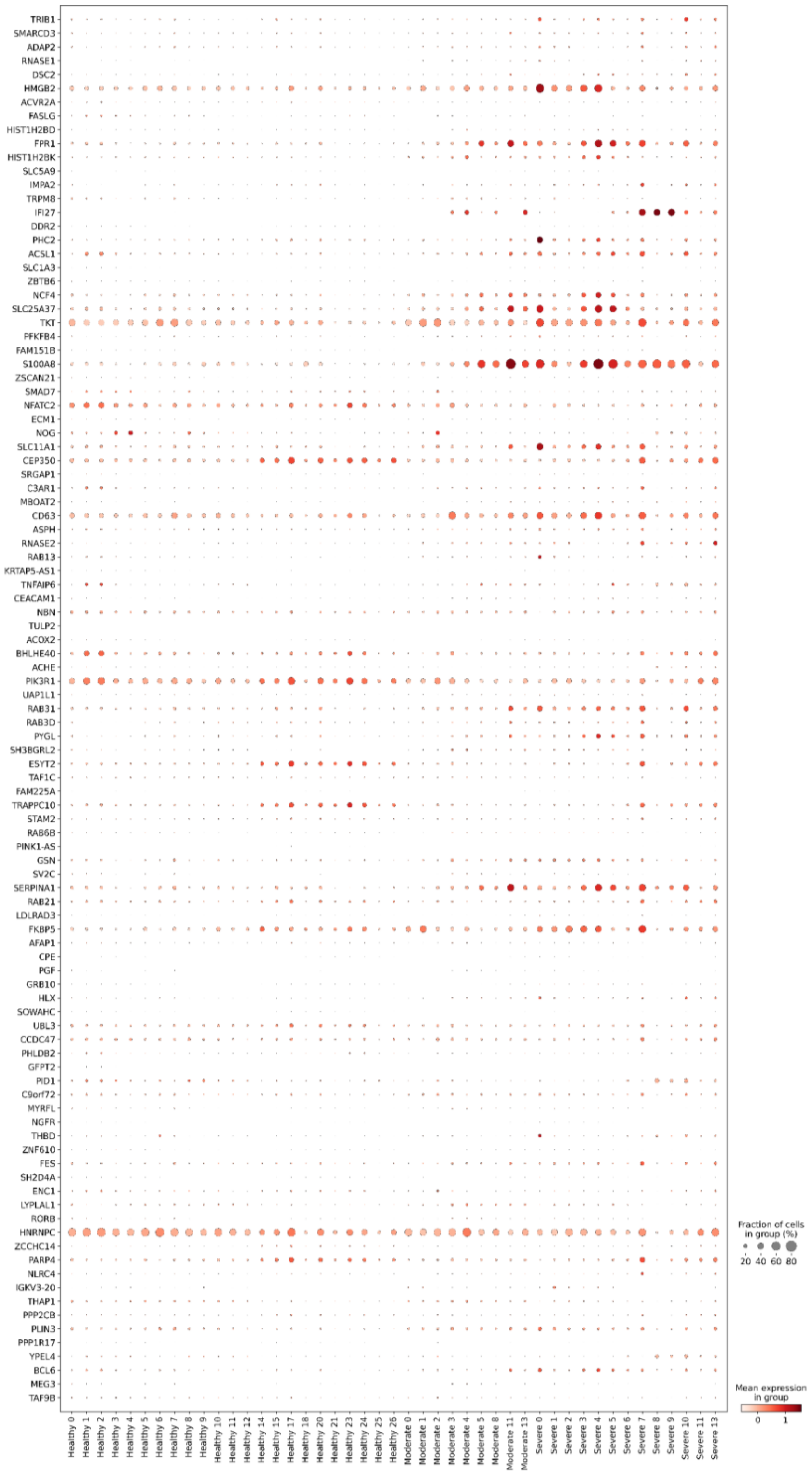


Figure S3. Dot plot depicting the average mean expression (indicated by color) and the percentage of expressing cells (indicated by size) of top 100 genes selected by Random Forests algorithm.
